# Neuropsychiatric, cognitive and brain morphology characteristics of conversion from Mild Cognitive Impairment to Alzheimer’s Disease

**DOI:** 10.1101/2021.12.01.21267124

**Authors:** Ronat Lucas, Hanganu Alexandru, for the ADNI group

## Abstract

The impact of neuropsychiatric symptoms (NPS) on cognitive performance has been extensively reported, and this impact was better defined in the aging population. Yet a potential impact of NPS on brain morphology, cognitive performance and interactions between them in a longitudinal setting, as well as the potential of using these values as prediction of conversion – have remained questionable. We studied 156 participants with mild cognitive impairment (MCI) from the *Alzheimer’s Disease Neuroimaging Initiative* database who maintained the same level of cognitive performance after a 4-year follow-up and compared them to 119 MCI participants who converted to dementia. Additionally, we assessed the same analysis in 170 healthy controls who remained healthy at follow-up. Compared to 15 controls who converted to MCI. Their neuropsychological, neuropsychiatric, and brain morphology data underwent statistical analyses of 1) baseline comparison between the groups; (2) analysis of covariance model controlling for age, sex, education, and MMSE score, to specify the cognitive performance and brain structures that distinguish the two subgroups, and 3) used the significant ANCOVA variables to construct a binary logistic regression model that generates a probability equation for a given individual to convert to a lower cognitive performance state.

Results showed that MCI who converted to AD in comparison to those who did not convert, exhibited a higher NPS prevalence, a lower cognitive performance and a higher number of involved brain structures. Furthermore, agitation, memory and the volumes of inferior temporal, hippocampal and amygdala sizes were significant predictors of MCI to AD conversion.

## Clinical states of age-related cognitive decline

Current clinical evaluations of cognitive decline include only two stages: (a) the mild cognitive impairment (MCI) and (b) dementia (or major cognitive impairment). These stages are based on cognitive markers (APA 2013; Jak et al. 2009; Albert et al. 2011) that are evaluated with comprehensive neuropsychological assessments. One of the leading clinical presentations of dementia is the Alzheimer’s disease (AD) type (Heidebrink 2002). An MCI level that would lead to AD has been characterized by either subjective concern about a change in cognition, or a lower performance in one or more cognitive domains in comparison to those expected for the patient’s age and educational background, without significant impairment in social or occupational functioning (Albert et al. 2011). The condition of reduced cognitive performance induces the existence of MCI single or multi-domain (Csukly et al. 2016; Petersen & Negash 2008).

On the other hand, in the demented state, cognitive deficits are sufficiently extensive that the individual is no longer able to carry out his or her daily life tasks alone or without supervision. These cognitive stages are frequently accompanied by psychological suffering for patients, relatives and caregivers (Bednarek et al. 2016; Schulz et al. 2008) as well as psychological and behavioral disturbances called neuropsychiatric symptoms (NPS)(Cummings et al. 1994).

## Neuropsychiatric symptoms in cognitive decline

Most of NPS are clearly observed in dementia (Cummings et al. 1994), but they also occur in the MCI stage (Rosenberg et al. 2013) and can be present in cognitively healthy individuals (Mortby et al. 2018). NPS presence was shown to increase the risk of AD in MCI (Teng et al. 2007). Indeed, these NPS are found in the cognitively normal (CN) population (Geda et al. 2004; Geda et al. 2008; Mortby et al. 2018), and their prevalence increases with the advancement of clinical stages: it is higher in the MCI population and even higher in the AD population (Geda et al. 2004). Also, they may increase the likelihood of MCI progressing into AD (Teng et al. 2007) and thus increase the likelihood of developing dementia (Rosenberg et al. 2013). These include depression (Lee et al. 2012; Steenland et al. 2012), apathy (van Dalen et al. 2018) and anxiety (Rozzini et al. 2009; Gallagher et al. 2011) (cited by Ma 2020), but the latter is more controversial (Robert et al. 2008; Devier et al. 2009). However, the impact of depression on cognitive decline is greater in MCI than in AD (Lee et al. 2019).

In addition, several studies have looked at longitudinal follow-ups of participants and patients with NPS. For example, the study by Moon et al (2017) confirms a greater progression from MCI to AD in patients with depressive symptoms according to the amyloid status of MCI patients: the study is based on the analysis of longitudinal ADNI data and shows, in MCI patients with amyloid-positive amyloid and depression, a higher rate of AD conversion than patients without depression. In addition, cognitive decline is accelerated over the 2-year follow-up period. Also based on the ADNI database, Zahodne et al. (2013) studied the atrophy pathways of MCI subjects with and without depression and apathy on a longitudinal level. Their results show that depression is associated with greater baseline entorhinal atrophy and accelerated anterior cingulate atrophy. To our knowledge, fewer studies have looked at the factors of conversion from normal cognition to MCI and the course of cognitive decline in healthy individuals. However, these studies were able to highlight that healthy individuals with mild behavioral impairment exhibited greater attentional and working memory decline after one year of follow-up (Creese et al. 2019). Also, the presence of NPS, including depression, apathy, and anxiety, is also associated with faster global and domain-specific decline (Burhanullah et al. 2020; Krell-Roesch et al. 2021). In addition, MRI data were also exploited as predictors of conversion from MCI to AD. Thus, it has been shown that MCI that convert to AD have reduced volumes in the medial temporal lobe (hippocampus, amygdala, and entorhinal cortex), the insular, posterior cingulate, precuneus and orbitofrontal cortex (Davatzikos et al. 2011; deToledo-Morrell et al. 2004; Risacher et al. 2009). However, these data do not appear to have been addressed in the conversion from CN to MCI. This shows the importance of screening for NPS and to more investigate MRI in CN subjects.

Fort his study, we hypothesized that (1) participants who convert to a lower cognitive performance state would exhibit increased variation in NPS; (2) these variations would be associated with brain morphology and cognitive performance, and (3) these correlations can be used to predict the conversion.

## Materials and Methods

### Participants

275 patients with MCI and 185 cognitively normal participants (CN) from the ADNI database were extracted. Data used in the preparation of this article were obtained from the ADNI database (adni.loni.usc.edu). The ADNI, launched in 2003 and led by Principal Investigator Michael W. Weiner, MD, has for main objective to understand the progression of MCI and early AD by combining imaging, biological and neuropsychological data (Mueller et al. 2005; Shaw et al. 2007) (http://www.adni-info.org/). Entry criteria for patients with amnestic MCI include a Mini-Mental State Examination score of 24 to 30 and a Memory Box score of at least 0.5, whereas other details on the ADNI cohort can be found online. All patients with AD met National Institute of Neurological and Communication Disorders/Alzheimer’s Disease and Related Disorders Association criteria for probable AD with a Mini-Mental State Examination score between 20 and 26, a global Clinical Dementia Rating of 0.5 or 1, a sum-of-boxes Clinical Dementia Rating of 1.0 to 9.0, and, therefore, are only mildly impaired. Exclusion criteria included any serious neurological disease other than possible AD, any history of brain lesions or head trauma, or psychoactive medication use (including antidepressants, neuroleptics, chronic anxiolytics, or sedative hypnotics).

Retained participants had completed a neuropsychiatric examination, a comprehensive neuropsychological assessment and a 3 Tesla MRI. The clinical status was available until 4-years follow-up for all participants. Based on the clinical stage at follow-up, four groups were created in order to distinguish participants who converted to a worse cognitive performance compared to those that maintained their previous cognitive level. Our groups consisted of: 156 MCI remained MCI at follow-up (MCI-non-converted), 119 MCI participants that converted to AD (MCI-converted), 170 CN both at baseline and at follow-up (CN-non-converted) and 15 CN that converted to MCI (CN-converted) (Table 1).

**Table 1:**
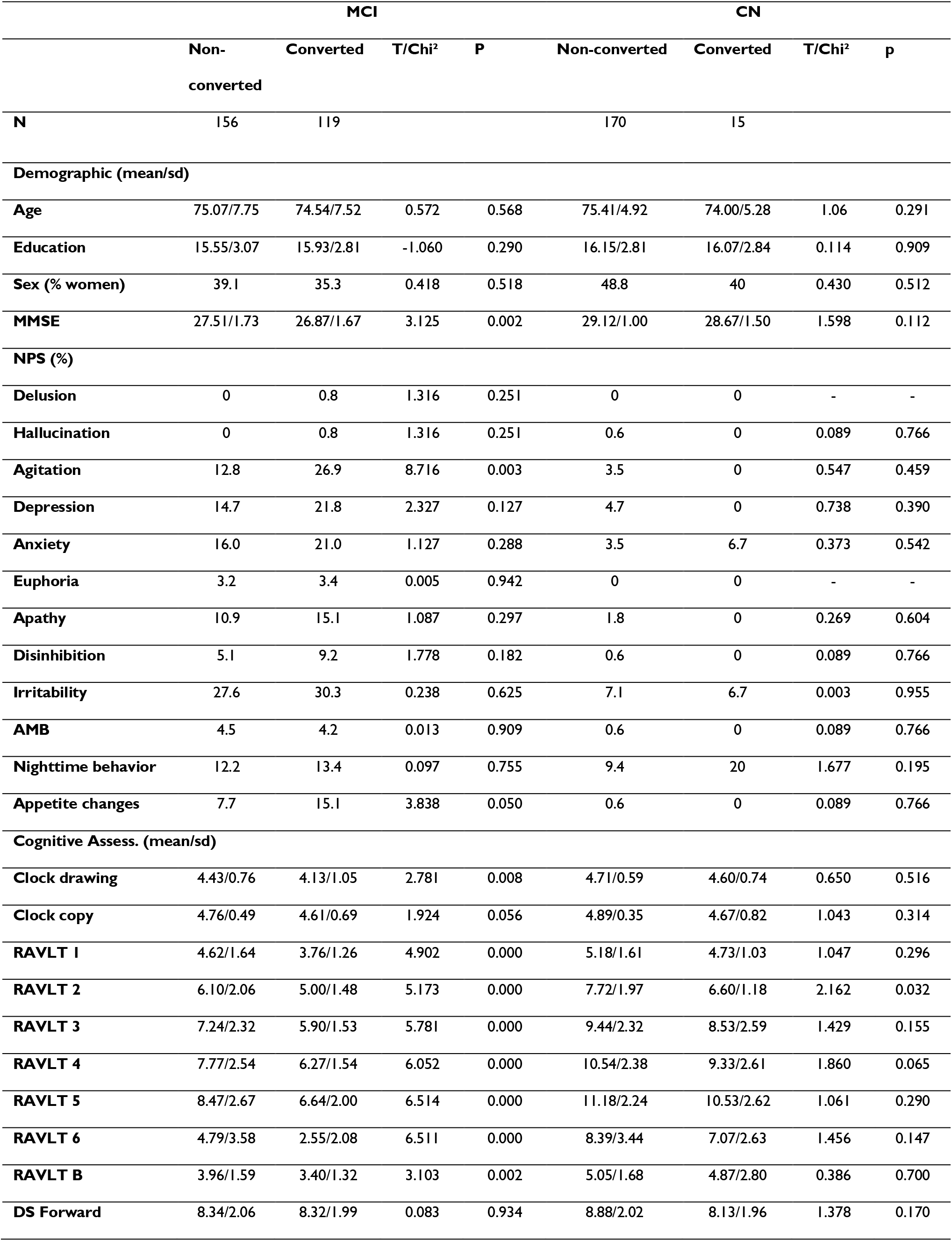
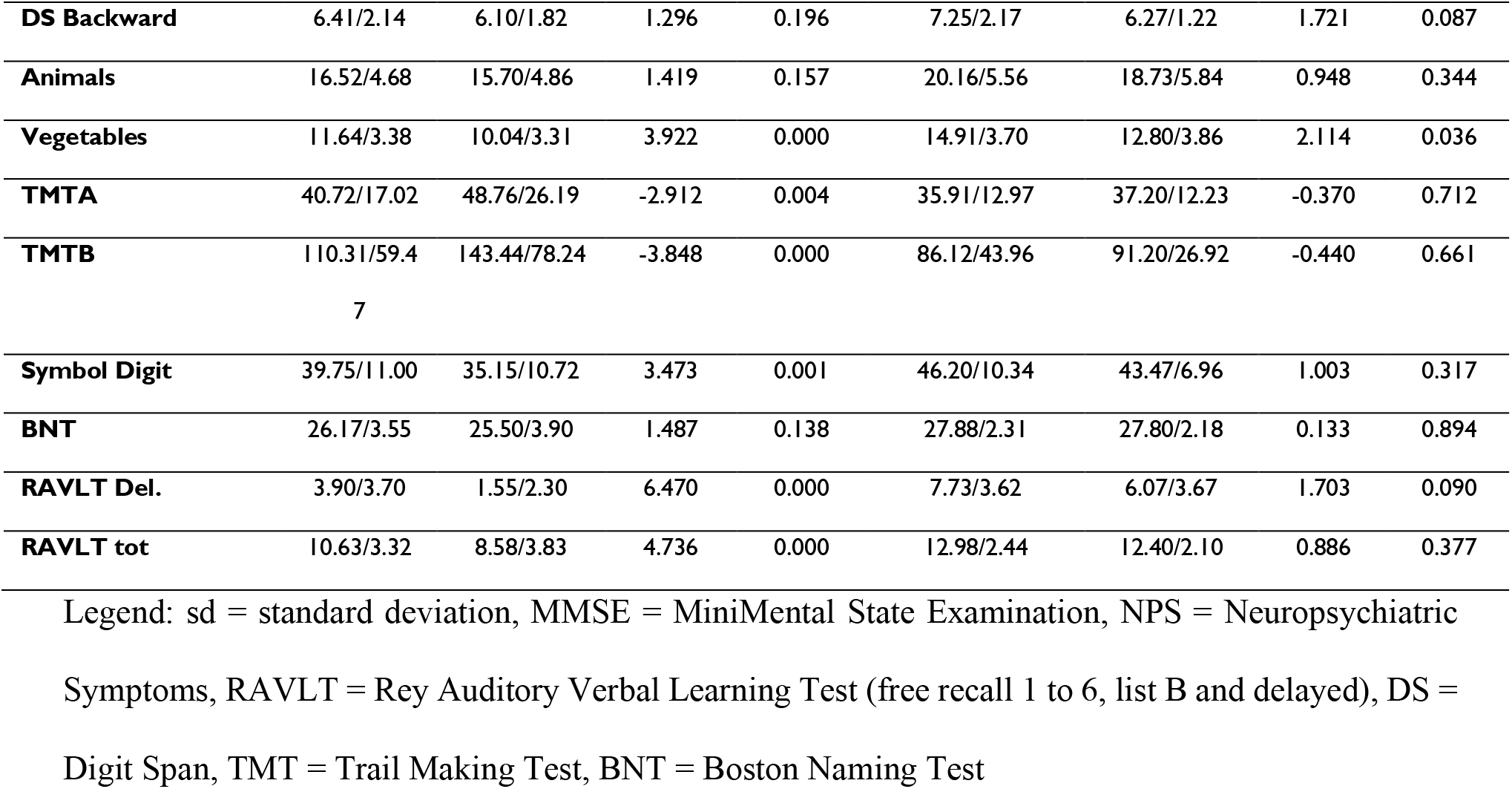
Demographic, neuropsychiatric and neuropsychological characteristics for MCI and CN groups (non-converted and converted).

### Data acquisition and processing

The neuropsychiatric changes were evaluated using the Neuropsychiatric Inventory (Cummings et al. 1994). The inventory consists of the evaluation of the presence, severity and frequency of 12 neuropsychiatric symptoms (NPS): delusions, hallucinations, agitation/aggressiveness, depression, anxiety, euphoria, apathy, disinhibition, irritability, aberrant motor behaviors, nighttime behaviors and appetite changes. We included the evaluations performed by the participants’ relatives and only the prevalence of each NPS is considered.

Neuropsychological assessment was based on the tests assessing: (1) anterograde verbal memory (Rey Auditory Verbal Learning Test - RAVLT), (2) focused attention (Trail Making Test A - TMTA), (2) processing speed (Wechsler Adult Intelligence Scale Code subtest), (3) mental flexibility (Trail Making Test B - TMTB), (4) visuoconstructive planning (clock test), (5) working memory (digit span), (6) semantic lexical evocation (animal and vegetable fluency) and (7) oral naming (Boston Naming Test - BNT). Moreover, the Mini-Mental State Examination score was used as a demographic factor of global cognitive efficiency.

MRI structural images were processed with FreeSurfer 7.1.1 software, on Linux Centos 7 on ComputeCanada environment, cluster Cedar and managed with our in-house pipeline (github.com/alexhanganu/nimb) that allowed automated exclusion of post-processed data with errors as well as extraction of statistical data, diminishing potential human error. Cortical thickness parameter was extracted based on the Destrieux et al. atlas (2010) while subcortical volumes were extracted for all regions as well as sub-regional based on the corresponding atlases; for the thalamus (Iglésias et al. 2018), amygdalas (Saygin et al. 2017), and hippocampi (Iglésias et al. 2015). The volumes of subcortical structures were corrected with the estimated Total Intracranial Volume (eTIV) (Sanfilipo et al. 2002).

### Statistical analysis

The statistical analyses are based on the methodology of Orso et al. (2020). For this study, the data were analyzed using SPSS version 26.0 software. Descriptive analyses verified the similarity of the groups (MCI-converted vs. MCI-non-converted; CN-converted vs. CN-non-converted) in terms of age, years of education, MMSE score and sex distribution (respectively mean comparisons by Student test and contingency Chi^2^ analysis). Statistical analysis consisted of three steps. (I) First, the groups were compared based on (i) means of cognitive performances (Two sample t-tests), (ii) prevalence of NPS (Chi^2^ tests) and (iii) means of neuroimaging structure sizes (Student t-tests of cortical thickness and subcortical volumes). (II) Features that were shown to be significant in the first three comparisons, were included in the Analysis of Covariance Model with age, sex, years of education and MMSE score as covariates. (III) Finally, features that were deemed significant in the ANCOVA model were imputed in a two binary logistic regression model to generate probability equations for AD and MCI conversion based on neuropsychiatric, cognitive and neuroimaging data.

## Results

### Demographical, neuropsychiatric and neuropsychological differences

At baseline, in comparison to MCI-non-converted, the MCI-converted group had a lower MMSE score, worse performance on some cognitive tests (clock test, RAVLT immediate recall A and B, RAVLT delayed recall, semantic lexical evocation for “vegetables”, TMT A and B, WAIS code) and a significantly higher prevalence of agitation and appetite changes (Table 1).

On the other hand, the CN-converted and CN-non-converted groups showed similar results regarding age, years of education, gender distribution, MMSE score as well as distributions of neuropsychiatric symptoms. Several significant differences were depicted in the cognitive performance at baseline, with the CN-converted group having a worse performance in comparison to CN-non-converted on RAVLT 2nd recall, digit span backward and semantic lexical evocation of “vegetables”.

### Brain morphology differences

In terms of brain morphology, the MCI-converted group also showed multiple significant difference in comparison to the MCI-non-converted one. Significant changes were depicted in all brain lobes both on the cortical level in gyri and sulci as well as regarding the volumes of subcortical structures, notably the volumes of hippocampus, amygdala and thalamus subregions (Supplementary table 1).

By contrast, the CN-converted group, when compared to the CN-non-converted one, showed a smaller number of brain morphological changes. Specific differences were found in the frontal inferior orbital gyrus and suborbital sulcus, cingulate ventral posterior gyrus, temporal pole and temporal middle gyrus. CN-converted also had reduced volumes in some hippocampal subregions (tails, subiculum, presubiculum, molecular layer, CA1 body), amygdala and thalamic subregions (central lateral, pulvinar anterior nuclei, pulvinar medial nuclei, paratenial nucleus) (Supplementary table 2).

### ANCOVA model

Concerning cognitive performance, in MCI to AD conversion, ANCOVA shows a significant lower performance in MCI-converted for every score of the RAVLT, and on semantic lexical evocation “vegetables” than in MCI-non-converted and lower performance in patients with appetitive changes for the 2nd free recall of the RAVLT. Only one interaction was found with agitation on the recall of the B-list of the RAVLT (non-converted with agitation perform better than those without while the opposite trend is present in converted). By contrast, the CN-converted group, when compared to the CN-non-converted one, exhibited significative lower performance on the 2nd recall of the RAVLT, and in semantic lexical evocation of “vegetables” (Supplementary table 4).

Regarding brain structures, most of the structures (all lobes, hippocampi and amygdala) were smaller in MCI-converted in relation to MCI-non-converted whereas agitation was featured by greater cortical thicknesses (frontal, parietal, occipital, temporal) and subcortical volumes (hippocampus and amygdala). In contrast, patients with appetite changes had precentral thinning and larger hippocampus and amygdala volumes than those without (Supplementary Table 3). Also, there was an interaction effect between conversion to AD and agitation on the occipital, cingulate, parietal and precentral structures. Agitation in MCI-converted patients is featured by: larger structure in occipital lobe and smaller left subcallosal gyrus and parieto-occipital sulcus; whereas a greater inferior precentral thinning in MCI-non-converted than converted was found. Interaction between conversion and appetite changes show that converted patients with appetite changes had smaller for occipital and parietal structures than patients without. The reverse pattern was found in non-converted patients (Supplementary table 3). In CN-converted, the previous structures (Supplementary table 2) remained significantly smaller than in CN-non-converted, except for the central lateral and paratenial thalamic nuclei (Supplementary table 4).

### Prediction of AD from MCI and MCI from CN based on logistic regression

Binary logistic regression models (Tables 2, 3) based on the significant results of the ANCOVA models provided probabilistic prediction equations for conversion of CN participants to MCI and MCI patients to AD.

**Table 2:** Binary Logistic Regression for the conversion from MCI to AD

| **Variables** | **B** | **Sig.** |
| --- | --- | --- |
| **Age** | -0,070 | 0,003 |
| **Free recall 1 of RAVLT** | -0,275 | 0,024 |
| **Free recall 5 of RAVLT** | -0,233 | 0,003 |
| **Agitation** | 0,765 | 0,049 |
| **Parietal precuneus gyrus left** | -2,425 | 0,012 |
| **Temporal inferior sulcus right** | -4,026 | 0,000 |
| **Hippocampus tail right** | -10665,677 | 0,004 |
| **Hippocampus fimbria right** | -26915,115 | 0,044 |
| **Amygdala Accessory Basal nucleus right** | -18399,454 | 0,024 |
| **Constante** | 28,139 | 0,000 |

**Table 3:**
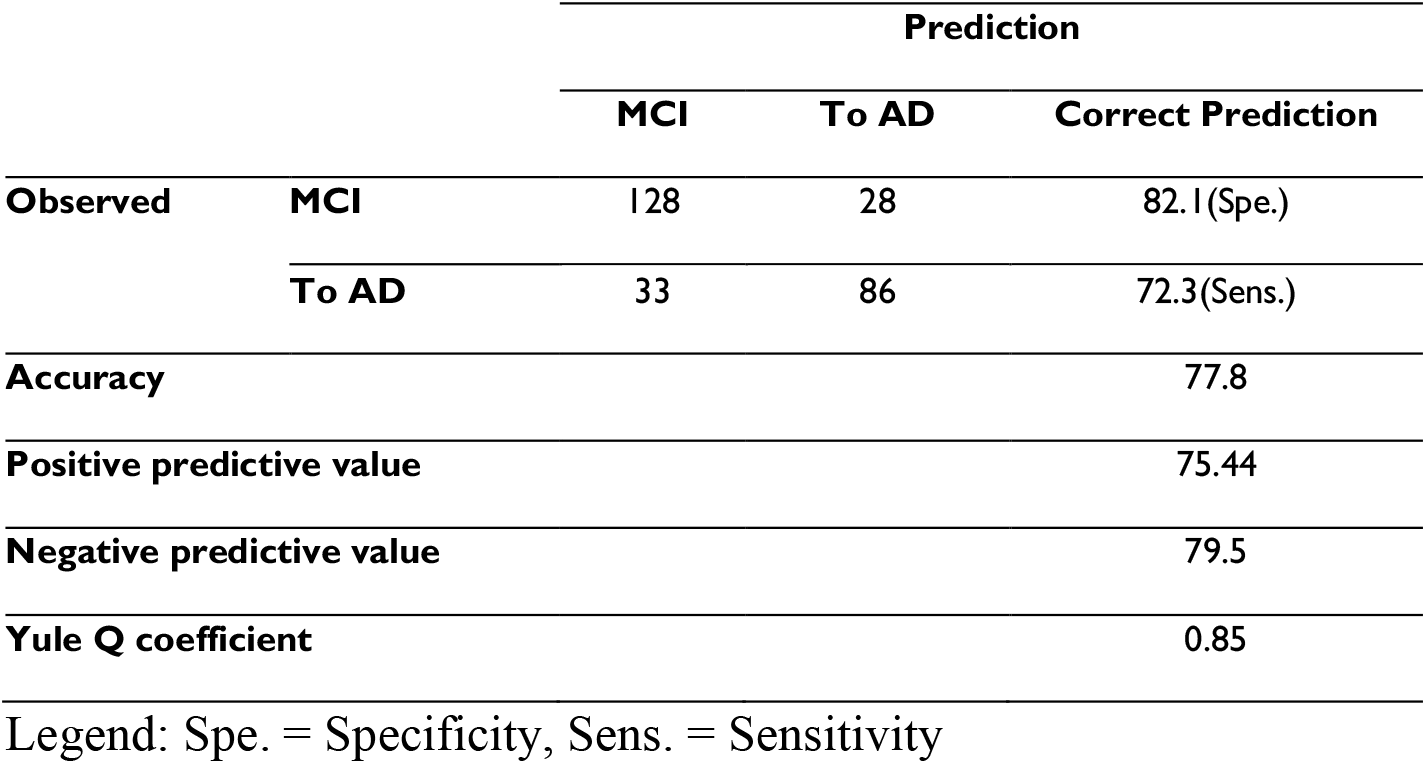
Psychometric characteristics for regression model of the conversion from MCI to AD

The probability equation for an MCI participant to convert to AD is as follows: 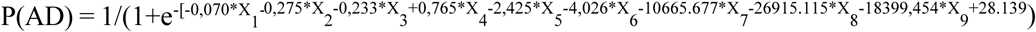 where e = 2,71828 (the base of natural logarithm), X_1_ = Age, X_2_ = the RAVLT 1^st^ immediate recall score, X_3_ = the RAVLT 5^th^ immediate recall score, X_4_ = the presence of agitation, X_5_ = the left precuneus gyrus thickness, X_6_ = the right inferior temporal sulcus thickness, X_7_ = the right hippocampal tail volume, X_8_ = the left hippocampal fimbria volume, X_9_ = the left amygdala accessory basal nucleus volume and 28.139 is the model’s constant. The model had a sensitivity of 72.3%, specificity of 82.1% (Tables 3), positive and negative predictive value of 75.44% and 79.5% and Yule Q coefficient ([true positives*true negatives - false positives*false negatives] / [true positives*true negatives + false positives*false negatives]) was 0.85, indicating a very strong link between the diagnosis and the clinical characteristics.

Then, the probability equation for a CN participant to convert to MCI is as follows: 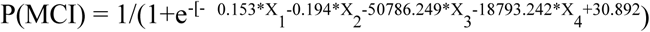 where e = 2,71828 (the base of natural logarithm), X_1_ = Age, X_2_ = the semantic lexical evocation for “vegetables” performance, X_3_ = the right subiculum body volume, X_4_ = the left medial pulvinar thalamic nucleus volume, and 30.892 is the model’s constant. The models were characterized by a sensitivity and specificity of 6.7% and 99.4% a (Table 5), positive and negative predictive values were 50% and 92.35%. Yule Q coefficient was 0.85 indicating a very strong link between the diagnosis and the clinical characteristics (Tables 4, 5).

**Table 4:** Variables significantly involved in predicting conversion from CN to MCI

| **Variables** | **B** | **Sig.** |
| --- | --- | --- |
| **Age** | -0,153 | 0,027 |
| **Semantic lexical evocation « vegetables »** | -0,194 | 0,036 |
| **Hippocampus, subiculum body, right** | -50786,249 | 0,011 |
| **Thalamus, pulvinar medial, left** | -18793,242 | 0,001 |
| **Constante** | 30,892 | 0,000 |

Legend: B = Coefficient from each significative variable, Sig. = Significancy.

**Table 5:**
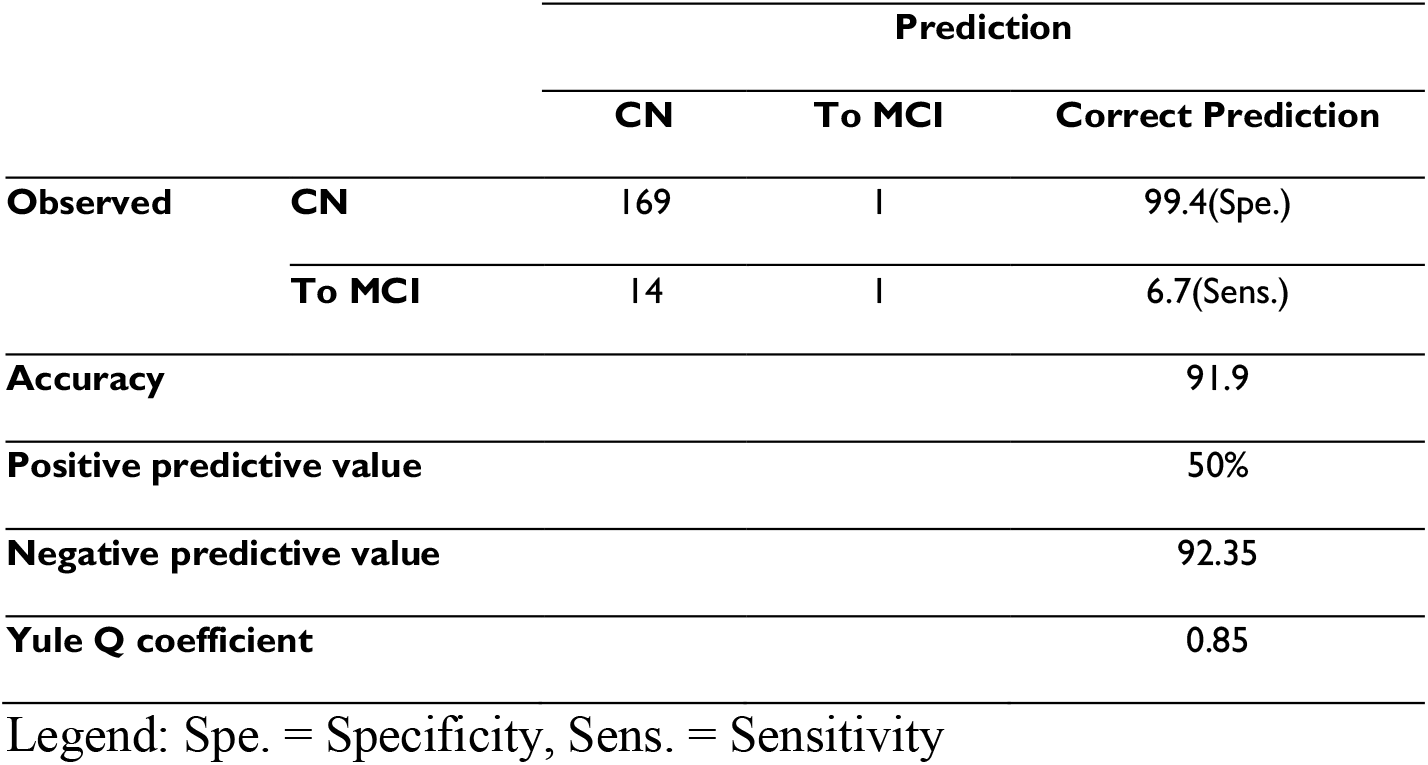
Psychometric characteristics for regression model of the conversion from CN to MCI

## Discussion

Our results show that (1) participants with MCI and CN who maintain their cognitive performance at the 4 years follow-up, tend to exhibited (i) a lower NPS prevalence, (ii) a higher cognitive performance and (iii) a lower number of involved brain structures; (2) from all NPS, only the presence of agitation might have the highest potential of predicting MCI participants who might convert in AD over 4 years;

(2) from all cognitive performance tests, only poorer mnesic performances seems to predict MCIs who convert to AD over 4 years, and only language performance might predict CN who convert to MCI over 4 years; (3) brain regions that seem to have the highest relevance in predicting conversion over 4 years, seem to be the hippocampus, amygdala, temporal inferior and parietal precuneus in the case of MCI participants, and hippocampus and thalamus in the case of Cn participants.

The involvement of agitation in our prediction model and absence of other NPS is of particular interest. Indeed, agitation along with appetite changes first appeared to have a significant higher prevalence in the MCI-converted group. Previous studies also outlined this potential trend by reporting agitation a precursor to future AD development (Gallagher et al. 2017; Lü et al. 2021; Rosenberg et al. 2013; Teng et al. 2007) and a sign in MCI participants that would correspond to an early AD diagnosis (Geda et al. 2014). Yet no specific link was reported regarding appetite changes. Furthermore, the involvement of other NPS has been reported: depression, anxiety, apathy, irritability, psychotic symptoms. Though in our model these NPS did not survive the significant threshold for the prevalence, nor did they appear in the prediction model, they did show a non-significant up to double increase in prevalence. This inconsistency might be due to (i) a difference in the NPS included in the model, (ii) significant difference in the number of participants as well as (iii) too high constraints used in our study. We would also outline the potential difference in the results regarding the involvement of NPS in the conversion of CN to MCI. Whereas several studies have been able to describe that the presence of NPS in CN increased the risk of conversion to MCI (Mortby et al. 2018; Lee et al. 2012, Steenland et al. 2012; van Dalen et al. 2018; Gallagher et al. 2011), our results found only an increased prevalance of anxiety and nighttime behavior changes in the CN-converted group in comparison to CN-non-converted, and yet, this increased prevalence didn’t seem to have a high potential for prediction. Nevertheless, this particular result we report only as a trend, due to the low number of CN-converted participants.

Cognitively, our results show lower verbal mnesic performance and semantic lexical evocation in MCI-converted vs. MCI-non-converted. Interestingly, we do not find executive weaknesses in MCI-converted although these deficits are frequent in MCI in relation to NPS (Rosenberg et al. 2011) as well as in AD (Grober et al. 2009). In addition, mnesic difficulties occur much earlier than the diagnosis of AD in comparison to executive difficulties (Grober et al. 2009). The CN-converted vs. CN-non-converted showed worse verbal mnesic, working memory and semantic lexical evocation performance. These performances cannot be considered as deficits in CN participants (because their performance remains within the populational norms established by the standardization of the tests), but they can probably be cognitive fragilities and signs of a beginning of cognitive decline, potentially in line with subjective cognitive complaints, not objectified by the neuropsychological tests (Edmonds et al. 2014; Mitchell et al. 2014; van Oijen et al. 2007). As such, only memory and semantic lexical evocation remained significantly involved in predicting conversion to AD and to MCI in our logistic regression models.

Regarding the significant role of memory performance, this is in line with the results of Baerresen et al. (2015). The use of this type of model, with predictive purposes, is more frequent in recent years and could be applied in individuals with non-amnestic MCI (San Lee et al. 2018), MCI due to Parkinson’s disease (Chen et al. 2020) or even multiple sclerosis (Eijlers et al. 2018). Unfortunately, these studies do not systematically mention the reliability criteria of their models.

Concerning brain difference between MCI-converted and MCI-non-converted, brain characteristics were broader and involve cortical structures of all lobes and subcortical regions of the hippocampus and amygdala. This suggests that diffuse cerebral frailties may already be present at the MCI stage, prior to the diagnosis of AD. However, only left precuneus, right temporal inferior, right hippocampus and right amygdala remained significantly predictive of conversion in the logistic regression model. Other studies have also shown cortical thinning of several lobes in MCI patients and even more so in AD, with a more important involvement of the left hemisphere (Singh et al. 2006; Wang et al. 2009). This asymmetry is also found in our data.

When analyzing specific brain changes from the perspective of involved NPS (agitation and appetite changes) and their potential to influence the brain in MCI-converted participants, previous studies showed that agitation was characterized by insular, superior frontal, middle, orbital, parieto-occipital, hippocampal, and amygdala atrophies in MCI (Hsu et al. 2015; Hu et al. 2015; Trzepacz et al.., 2013). Furthermore, these atrophies were broadly similar between MCI and AD patients. Our data comparing the effect of agitation in converters and non-converters instead showed occipital, cingulate, precentral, superior temporal, parasubiculum, and amygdala features. Note that the impact of agitation in non-converters was characterized mainly by reductions in structure size, and by increases in size in converters. This might suggest that the underlying physiological processes are not the same (e.g. atrophy vs. compensation or inflammation). According to Bateman et al. (2020), pro-inflammatory processes have a positive correlation with agitation in AD while anti-inflammatory processes have a negative correlation with agitation. Since the increase in brain structures here is only observed in our converted group, we should look at the age of the agitation. It could be assumed that in the converted group, agitation is older and could have allowed the development of inflammatory processes.

Interestingly, appetite changes seem more prevalent in MCI converted in AD than in non-converted but them were not retained by regression models as significant factor who predict the conversion. Frequently, these behavioral changes are mainly associated with posterior structures. Particularly, them have been described in patients with posterior cortical atrophy but associated with posterior structures also in typical AD (Isella et al. 2015), indicating that posterior brain damage is not specific to these disorders. Overall, these are understudied disorders and often dependent on other NPS such as anxiety or depression (Ismail et al. 2008; Suma et al. 2018). This makes these disorders more complicated to study, especially on a neuroanatomical level.

In the CN to MCI conversion, the poorer performance in memory suggested that the brain structures involved in memory would be reduced in people who convert to MCI. However, the cortical and subcortical structures involved appear to be broader and involved in emotional (cingular, amygdala, frontal orbital), memory (temporal, hippocampus) and multimodal functions (thalamus). According to regression model, smaller volumes in the right hippocampus and the left thalamus predicted better the conversion to MCI. Previous studies that have looked at brain differences have focused on comparisons between CN individuals and individuals with MCI. These studies showed reductions in hippocampal, entorhinal and parahippocampal cortex volumes (Jessen et al. 2006; Saykin et al. 2006). In their study Gifford et al. (2015) compared patients with MCI with vs. without subjective memory complaints (SMC). Their results show lower performance in immediate and delayed episodic memory assessed on a serial list learning task in individuals with subjective memory complaints, but no difference in volumes and thicknesses of brain regions of interest (lobar volumes: frontal, parietal, temporal, cingulate; and specific medial temporal lobe structures: hippocampal volume, entorhinal cortex thickness, parahippocampal gyrus thickness). In addition, Schultz et al. (2015) studied cognitively normal subjects with vs. without SMC. Their results showed that, compared with individuals without SMC, those with SMC had significant cortical thinning in the entorhinal, fusiform, posterior cingulate, and inferior parietal cortices and significantly reduced amygdala volume. Similarly, those with SMC had significantly lower test scores on measures of Immediate Memory, Verbal Learning & Memory, and Verbal Ability (Schultz et al. 2015). One explanation from Gifford et al (2015) about their absence of brain differences between MCI with vs without SMC is that null finding could suggest that MCI individuals who report a memory change have medial temporal and global atrophy comparable to MCI individuals who deny any memory changes, making detection of any between-group differences difficult. Other studies, conducted in a large community-based sample, SMC were associated with cross-sectional decrements in hippocampal, parahippocampal, and amygdala volumes (Stewart et al. 2008) and longitudinal hippocampal volume loss 4 years later (Stewart et al. 2011).

To our knowledge, most studies have focused on regions of interest known to be involved in Alzheimer’s disease, whereas our study looked at the entire cortex and subcortical structures. Furthermore, our results may suggest brain changes that precede medial temporal damage, which is usually considered as an anatomical precursor of cognitive decline due to Alzheimer’s disease.

Because the risk of developing MCI was dependent on certain demographic data, we chose to include them in the regression model, whereas these variables were controlled in the ANCOVA model to isolate differences related to cognitive performance and brain structure size. Other analyses, on other databases, should also consider the systematic presence of SMC in CN individuals, as well as the presence of symptoms related to awareness of changes and difficulties (anosognosia and/or anosodiaphoria). Alternatively, if our results do not show neuropsychiatric differences in CN-converted, this may suggest the existence of “subjective behavioral complaints” that would precede the objectification of a mild behavioral disorder, as described by Ismail et al. (2016, 2017), in analogy to the stages of cognitive decline model.

One limitation of this study concerns the small proportion of CN individuals who convert to MCI. Several factors could explain this low rate as well as the lack of NPS involvement in conversion to MCI: 1) many participants were not followed up due to dropout (significant attrition), 2) the presence of NPS increases the risk of stopping participation in database follow-up, this risk induces a selectivity bias of a lower proportion of longitudinal data in participants with NPS (Burke et al. 2019), 3) other factors increase attrition (older age, lower education and socioeconomic level)(Bhamra et al. 2008). This could explain the low sensitivity of our model. Furthermore, the presence of NPS is considered a risk factor for cognitive decline, not a necessary condition for decline. Thus, subgroups of individuals developing MCI without NPS can frequently be observed. Finally, the limited implication of neuromorphological characteristics may have reduced the possibility of distinguishing our two subgroups. Indeed, some data appear to support cerebral metabolic abnormalities in the preclinical stages of AD rather than structural abnormalities (Kljajevic et al. 2014; Ng et al. 2017).

## Conclusion

Research on the preclinical stages of AD is frequent and focuses on different diagnostic criteria and risk factors. As far as we know, our study is one of the first to apply these types of models with MCI and CN individuals using both neuropsychiatric, cognitive and neuromorphological data. Weproposed to distinguish MCI and CN participants who convert to AD and MCI, respectively, after 4 years of follow-up from the ADNI database. We were able to establish two predictive models to distinguish participants evolving to a more severe clinical stage. The conversion from MCI to AD was characterized by the presence of agitation, lower memory performance and smaller volumes of inferior temporal, hippocampal and amygdala brain structures, whereas the conversion from CN to MCI was characterized by lower performance on semantic evocation and smaller volumes of hippocampal and thalamic brain structures. The use of these analytical methods might be a good way to anticipate cognitive and brain declines.

## Data Availability

All data used are available online at http://adni.loni.usc.edu/

## Use of ADNI data

*Data collection and sharing for this project was funded by the Alzheimer’s Disease Neuroimaging Initiative (ADNI) (National Institutes of Health Grant U01 AG024904) and DOD ADNI (Department of Defense award number W81XWH-12-2-0012). ADNI is funded by the National Institute on Aging, the National Institute of Biomedical Imaging and Bioengineering, and through generous contributions from the following: AbbVie, Alzheimer’s Association; Alzheimer’s Drug Discovery Foundation; Araclon Biotech; BioClinica, Inc*.; *Biogen; Bristol-Myers Squibb Company; CereSpir, Inc*.; *Cogstate; Eisai Inc*.; *Elan Pharmaceuticals, Inc*.; *Eli Lilly and Company; EuroImmun; F. Hoffmann-La Roche Ltd and its affiliated company Genentech, Inc*.; *Fujirebio; GE Healthcare; IXICO Ltd*.; *Janssen Alzheimer Immunotherapy Research & Development, LLC*.; *Johnson & Johnson Pharmaceutical Research & Development LLC*.; *Lumosity; Lundbeck; Merck & Co*., *Inc*.; *Meso Scale Diagnostics, LLC*.; *NeuroRx Research; Neurotrack Technologies; Novartis Pharmaceuticals Corporation; Pfizer Inc*.; *Piramal Imaging; Servier; Takeda Pharmaceutical Company; and Transition Therapeutics. The Canadian Institutes of Health Research is providing funds to support ADNI clinical sites in Canada. Private sector contributions are facilitated by the Foundation for the National Institutes of Health (www.fnih.org). The grantee organization is the Northern California Institute for Research and Education, and the study is coordinated by the Alzheimer’s Therapeutic Research Institute at the University of Southern California. ADNI data are disseminated by the Laboratory for NeuroImaging at the University of Southern California*.

