## Supplemental tables for "Neuropsychiatric, cognitive and brain morphology characteristics of conversion from Mild Cognitive Impairment to Alzheimer’s Disease"

**Supplementary Table 1** : Structural differences between MCI-non-converted vs MCI-converted.

| Lobe | Structure | Left t | Left p | Right t | Right p |
|---|---|---|---|---|---|
| Frontal | superior gyrus | 2,465 | 0,014 | - | - |
| | superior sulcus | 2,544 | 0,011 | - | - |
| | middle gyrus | 3,337 | 0,001 | 2,491 | 0,013 |
| | middle sulcus | 2,705 | 0,007 | - | - |
| | inferior sulcus | 3,806 | 0,000 | 1,963 | 0,051 |
| | rectus gyrus | 1,964 | 0,051 | 2,630 | 0,009 |
| | precentral inferior part sulcus | 2,319 | 0,021 | 2,041 | 0,042 |
| | pole transverse gyrus and sulcus | 3,764 | 0,000 | 3,517 | 0,001 |
| | marginal gyrus and sulcus | 2,760 | 0,006 | 3,173 | 0,002 |
| | Inferioro opercularis gyrus | 3,049 | 0,003 | 2,511 | 0,013 |
| | inferior triangularis gyrus | - | - | 2,056 | 0,041 |
| | inferior orbital gyrus | - | - | 2,667 | 0,008 |
| | orbital H shaped sulcus | 1,987 | 0,048 | - | - |
| | orbital medial olfactive sulcus | - | - | 2,998 | 0,003 |
| Occipital | superior transversal sulcus | 3,024 | 0,003 | - | - |
| | middle gyrus | 3,470 | 0,001 | 2,268 | 0,024 |
| | middle lunatus sulcus | 2,075 | 0,039 | - | - |
| | inferior gyrus and sulcus | 2,087 | 0,038 | - | - |
| | anterior sulcus | 2,061 | 0,040 | - | - |
| | calcarine sulcus | 2,469 | 0,014 | 2,247 | 0,025 |
| | occipito-temporal lateral sulcus | 3,553 | 0,000 | 2,919 | 0,004 |
| | occipito-temporal medial lingual sulcus | 3,005 | 0,003 | 3,926 | 0,000 |
| Cingulate | anterior gyrus and sulcus | 2,003 | 0,046 | - | - |
| | posterior dorsal gyrus | 4,130 | 0,000 | 4,316 | 0,000 |
| | posterior ventral gyrus | 3,267 | 0,001 | 3,772 | 0,000 |
| | subcallosal gyrus | 2,810 | 0,005 | 2,646 | 0,009 |
| | mid Posterior gyrus and sulcus | - | - | 2,711 | 0,007 |
| Insula | circular superior Sulcus | - | - | 2,845 | 0,005 |
| | circular inferior Sulcus | 3,726 | 0,000 | 3,557 | 0,000 |
| | circular anterior Sulcus | 2,010 | 0,045 | - | - |
| | long insular gyrus and central sulcus | 4,136 | 0,000 | 3,681 | 0,000 |
| | short gyrus | 3,320 | 0,001 | - | - |
| Parietal | superior gyrus | 2,417 | 0,016 | 2,565 | 0,011 |
| | inferior angular gyrus | 4,439 | 0,000 | 3,807 | 0,000 |
| | inferior supramar gyrus | 4,561 | 0,000 | 2,880 | 0,004 |
| | precuneus gyrus | 4,092 | 0,000 | 3,807 | 0,000 |
| | Jensen sulcus | 3,535 | 0,000 | 2,933 | 0,004 |
| | intraparietal sulcus | 3,486 | 0,001 | 2,869 | 0,004 |
| | parieto-occipital sulcus | 3,026 | 0,003 | 2,613 | 0,009 |
| | subparietal sulcus | 2,664 | 0,008 | 3,506 | 0,001 |
| Temporal | superior lateral gyrus | 3,816 | 0,000 | 2,580 | 0,010 |
| | superior planum polare gyrus | 3,872 | 0,000 | 3,676 | 0,000 |
| | superior planum temporale gyrus | 3,427 | 0,001 | 2,832 | 0,005 |
| | superior sulcus | 4,647 | 0,000 | 4,287 | 0,000 |
| | middle gyrus | 4,677 | 0,000 | 5,275 | 0,000 |
| | inferior gyrus | 4,107 | 0,000 | 5,558 | 0,000 |
| | inferior sulcus | 4,527 | 0,000 | 5,873 | 0,000 |
| | lateral fusiform gyrus | 3,726 | 0,000 | 3,595 | 0,000 |
| | medial parahippocampal gyrus | 3,366 | 0,001 | 4,141 | 0,000 |
| | pole | 4,045 | 0,000 | 3,617 | 0,000 |
| | lateral fissure posterior | - | - | 3,116 | 0,002 |
| | collateral transverse anterior sulcus | - | - | 2,253 | 0,025 |
| | collateral transverse posterior sulcus | - | - | 2,759 | 0,006 |
| Hippocampus | whole | 5,708 | 0,000 | 5,815 | 0,000 |
| | whole head | 5,620 | 0,000 | 5,032 | 0,000 |
| | whole body | 5,137 | 0,000 | 5,659 | 0,000 |
| | presubiculum head | 4,870 | 0,000 | 4,649 | 0,000 |
| | presubiculum body | 2,910 | 0,004 | 3,787 | 0,000 |
| | subiculum head | 4,493 | 0,000 | 4,019 | 0,000 |
| | subiculum body | 4,262 | 0,000 | 5,168 | 0,000 |
| | ca1 head | 4,965 | 0,000 | 4,461 | 0,000 |
| | ca1 body | 3,083 | 0,002 | 3,600 | 0,000 |
| | ca3 head | 4,533 | 0,000 | 3,960 | 0,000 |
| | ca3 body | 4,544 | 0,000 | 4,800 | 0,000 |
| | ca4 head | 5,172 | 0,000 | 4,546 | 0,000 |
| | ca4 body | 4,498 | 0,000 | 5,053 | 0,000 |
| | hata | 4,450 | 0,000 | 4,097 | 0,000 |

| | | | | | |
|---|---|---|---|---|---|
| | fimbria | 4,243 | 0,000 | 3,284 | 0,001 |
| | molecular Layer head | 5,599 | 0,000 | 5,203 | 0,000 |
| | molecular Layer body | 5,122 | 0,000 | 6,025 | 0,000 |
| | gcmldg head | 5,271 | 0,000 | 4,713 | 0,000 |
| | gcmldg body | 4,654 | 0,000 | 5,019 | 0,000 |
| | tail | 4,303 | 0,000 | 5,562 | 0,000 |
| Amygdala | Lateral nucleus | 4,858 | 0,000 | 4,520 | 0,000 |
| | Basal nucleus | 5,445 | 0,000 | 5,435 | 0,000 |
| | Accessory Basal nucleus | 6,421 | 0,000 | 6,618 | 0,000 |
| | Anterior amygdaloid area | 4,823 | 0,000 | 3,891 | 0,000 |
| | Central nucleus | 6,772 | 0,000 | 5,876 | 0,000 |
| | Medial nucleus | 3,848 | 0,000 | 3,993 | 0,000 |
| | Cortical nucleus | 4,879 | 0,000 | 6,173 | 0,000 |
| | Cortico-amygdaloid transition | 4,346 | 0,000 | 4,399 | 0,000 |
| | Paralaminar nucleus | 3,107 | 0,002 | 3,653 | 0,000 |
| | Whole | 5,663 | 0,000 | 5,476 | 0,000 |
| Thalamus | LD | 2,241 | 0,026 | 2,493 | 0,013 |
| | LGN | 2,829 | 0,005 | 2,146 | 0,033 |
| | MDl | 2,332 | 0,020 | - | - |
| | MVRe | 2,227 | 0,027 | 2,520 | 0,012 |
| | PuA | 2,507 | 0,013 | - | - |
| | PuI | 2,539 | 0,012 | 2,010 | 0,045 |

Legend: gcmldg = granule cell layers of the dentate gyrus, CA = Cornu Ammonis, hata = Hippocampus amygdala transition area, LD = Laterodorsal, LGN = Lateral Geniculate Nucleus, MDl = Mediodorsal lateral, MVRe = Medial Ventral Reuniens, PuA = Pulvinar Anterior, PuI = Pulvinar Inferior.

**Supplementary Table 2**: Structural differences between CN-non-converted vs CN-converted.

| Lobe | Structure | Left t | Left p | Right t | Right p |
|---|---|---|---|---|---|
| Cingulate | Posterior ventral gyrus | - | - | 2.328 | 0.021 |
| Frontal | inferior Orbital gyrus | - | - | 2.027 | 0.044 |
| Temporal | middle gyrus | - | - | 1.973 | 0.050 |
| | pole gyrus | - | - | 2.001 | 0.047 |
| Hippocampus | whole | - | - | 2.032 | 0.044 |
| | whole body | - | - | 2.260 | 0.025 |
| | presubiculum body | - | - | 2.604 | 0.010 |
| | subiculum head | 2.349 | 0.020 | - | - |
| | subiculum body | - | - | 2.757 | 0.006 |
| | ca1 body | 2.167 | 0.032 | - | - |
| | molecular layer body | - | - | 2.472 | 0.014 |
| | tail | 2.039 | 0.043 | 2.287 | 0.023 |
| Amygdala | anterior amygdaloid area | 2.401 | 0.017 | - | - |
| | corticoamygdaloid transition | - | - | 2.109 | 0.036 |
| Thalamus | whole | 2.538 | 0.012 | - | - |
| | central lateral | 2.478 | 0.021 | - | - |
| | paratenial | - | - | 2.168 | 0.042 |
| | pulvinar Interior | 2.945 | 0.004 | - | - |
| | pulvinar medial | 3.364 | 0.001 | - | - |

Legend: t = statistical value from the Student T-test Model, Left = left hemisphere, right = right hemisphere, CA = Cornu Ammonis

**Supplementary Table 3**: Simple and interaction effects of group (MCI-converted vs MCI-non-converted) and NPS (Agitation and Appetite changes) on cognitive performance and brain structures from corrected ANCOVA model.

| | | Simple effects | | | Interaction effects | |
|---|---|---|---|---|---|---|
| | | Group | Agi | App | Group*Agi | Group*App |
| Domain | Variable | F-values | | | | |
| Cognitive | Free recall 1 RAVLT | 21,0*** | 0,6 | 1,2 | 0,2 | 3,7 |
| | Free recall 2 RAVLT | 8,7** | 1,5 | 4,0* | 0,0 | 0,1 |
| | Free recall 3 RAVLT | 11,7*** | 0,0 | 1,1 | 0,1 | 0,1 |
| | Free recall 4 RAVLT | 11,0*** | 0,0 | 0,1 | 0,0 | 0,1 |
| | Free recall 5 RAVLT | 12,5*** | 1,7 | 0,4 | 0,1 | 0,0 |
| | Free recall 6 RAVLT | 18,4*** | 1,0 | 0,0 | 0,0 | 2,5 |
| | Free recall B RAVLT | 10,7*** | 0,3 | 1,2 | 6,0* | 1,7 |
| | Semantic lexical evocation "Vegetables" | 9,4** | 0,4 | 0,3 | 0,6 | 0,5 |
| | Delayed free recall RAVLT | 16,3*** | 0,3 | 0,5 | 0,1 | 2,5 |
| | Total delayed recall RAVLT | 14,2*** | 0,2 | 0,1 | 0,1 | 1,7 |
| Frontal | rectus Gyrus Left | 0,6 | 1,8 | 0,1 | 0,2 | 0,4 |
| | rectus Gyrus Right | 5,6* | 0,1 | 0,0 | 2,1 | 1,1 |
| | precentral Inferior part Sulcus Left | 0,2 | 0,6 | 5,6* | 8,0** | 1,1 |
| | precentral Inferior part Sulcus Right | 0,0 | 0,7 | 1,2 | 4,3* | 0,3 |
| | Inferior Sulcus Left | 3,7 | 4,5* | 0,4 | 0,0 | 0,2 |
| | Inferior Triangularis Gyrus Right | 0,0 | 5,1* | 1,0 | 0,7 | 0,4 |
| Occipital | Superior transversal Sulcus Left | 5,0* | 7,6** | 0,4 | 2,2 | 1,4 |
| | middle Gyrus Left | 5,0* | 1,9 | 1,1 | 0,2 | 2,4 |
| | middle Lunatus Sulcus Left | 4,0* | 4,4* | 1,5 | 0,5 | 2,9 |
| | temporal Lateral Sulcus Left | 7,2** | 0,6 | 0,6 | 0,5 | 0,8 |
| | temporal Medial Lingual Sulcus Right | 11,8*** | 0,6 | 2,8 | 0,4 | 7,1** |
| | Inferior Gyrus and Sulcus Left | 0,3 | 0,2 | 0,0 | 6,2* | 0,1 |
| Cingulate | subcallosal Gyrus Left | 8,2** | 3,1 | 0,1 | 4,6* | 2,6 |
| | subcallosal Gyrus Right | 7,3** | 1,0 | 1,3 | 2,2 | 2,4 |
| Insula | circular Inferior Sulcus Left | 4,1* | 0,3 | 0,0 | 1,5 | 1,8 |
| Parietal | Superior Gyrus Left | 7,0** | 0,8 | 0,6 | 3,4 | 5,9* |
| | Superior Gyrus Right | 6,2* | 2,5 | 0,0 | 0,9 | 3,6 |
| | Inferior Angular Gyrus Left | 10,1** | 2,3 | 0,0 | 2,9 | 2,2 |
| | Inferior Angular Gyrus Right | 7,5** | 3,6 | 0,2 | 0,6 | 2,3 |
| | Inferior Superiorramar Gyrus Left | 7,7** | 0,0 | 0,8 | 1,5 | 2,4 |
| | Inferior Superiorramar Gyrus Right | 2,9 | 5,6* | 0,2 | 0,5 | 1,0 |
| | intraParietal Sulcus Left | 7,5** | 0,1 | 1,9 | 4,1* | 5,2* |
| | intraParietal Sulcus Right | 4,0* | 0,6 | 0,0 | 0,4 | 2,2 |
| | subparietal Sulcus Left | 8,3** | 3,7 | 0,4 | 1,3 | 7,3** |
| | subparietal Sulcus Right | 10,7*** | 1,4 | 2,8 | 0,5 | 6,9** |
| | parieto-occipital Sulcus Left | 5,9* | 1,5 | 0,4 | 4,0* | 2,1 |
| | parieto-occipital Sulcus Right | 8,9** | 2,0 | 1,7 | 0,1 | 8,9** |
| | precuneus Gyrus Left | 10,1** | 1,3 | 2,1 | 3,0 | 4,3* |
| | precuneus Gyrus Right | 10,5*** | 4,9* | 0,0 | 0,3 | 4,8* |
| Temporal | superior lateral Gyrus Left | 5,3* | 2,7 | 0,0 | 0,0 | 1,1 |
| | superior lateral Gyrus Right | 2,1 | 6,4* | 0,1 | 2,8 | 0,9 |
| | Superior sulcus Left | 7,7** | 1,6 | 0,6 | 0,2 | 1,1 |
| | superior sulcus Right | 4,3* | 0,8 | 0,2 | 1,1 | 0,6 |

| | | | | | | |
|---|---|---|---|---|---|---|
| | superior plan tempo Gyrus Right | 4,1* | 0,6 | 2,0 | 0,5 | 1,4 |
| | middle gyrus Left | 4,9* | 0,0 | 0,1 | 0,1 | 0,4 |
| | middle gyrus Right | 5,8* | 1,3 | 0,6 | 3,5 | 0,2 |
| | lateral fissure Posterior Right | 5,0* | 1,2 | 0,4 | 1,4 | 2,4 |
| | inferior gyrus Right | 7,0** | 0,1 | 1,6 | 1,5 | 0,3 |
| | Inferior sulcus Left | 4,3* | 0,4 | 1,8 | 1,0 | 0,0 |
| | inferior sulcus Right | 4,3* | 1,0 | 0,0 | 3,2 | 0,0 |
| **Hippocampus** | whole left | 8,1** | 1,3 | 5,1* | 0,1 | 2,1 |
| | whole right | 7,6** | 3,4 | 4,3* | 0,0 | 0,4 |
| | whole head left | 6,4* | 1,1 | 3,8 | 0,5 | 1,8 |
| | whole head right | 4,2* | 3,5 | 3,6 | 0,0 | 0,2 |
| | whole body left | 9,0** | 1,6 | 4,6* | 0,0 | 2,3 |
| | whole body right | 10,1** | 3,1 | 3,5 | 0,0 | 0,5 |
| | presubiculum head left | 5,7* | 1,9 | 3,6 | 1,2 | 1,0 |
| | presubiculum head right | 4,8* | 3,0 | 1,9 | 1,0 | 0,6 |
| | presubiculum body right | 7,5** | 1,1 | 0,3 | 0,0 | 1,8 |
| | subiculum head left | 5,8* | 1,2 | 2,1 | 0,3 | 1,5 |
| | subiculum body left | 7,8** | 1,4 | 3,4 | 0,2 | 3,3 |
| | subiculum body right | 12,6*** | 3,3 | 1,8 | 0,0 | 1,6 |
| | ca1 head left | 3,9* | 0,6 | 2,7 | 0,4 | 1,3 |
| | ca1 head right | 2,7 | 2,5 | 4,4* | 0,2 | 0,0 |
| | ca1 body right | 1,4 | 0,4 | 4,8* | 0,0 | 0,0 |
| | ca3 head left | 6,3* | 1,5 | 3,0 | 0,0 | 3,6 |
| | ca3 body left | 4,6* | 1,9 | 8,1** | 0,0 | 0,8 |
| | ca3 body right | 4,7* | 4,1* | 5,9* | 0,0 | 0,0 |
| | ca4 head left | 7,7** | 1,6 | 3,3 | 0,8 | 2,8 |
| | ca4 body right | 6,8** | 3,1 | 5,2* | 0,0 | 0,0 |
| | ca4 body left | 4,9* | 1,6 | 5,4* | 0,0 | 1,2 |
| | hata right | 3,9* | 1,6 | 0,0 | 1,8 | 0,0 |
| | fimbria left | 3,8 | 0,0 | 0,6 | 0,0 | 0,0 |
| | molecular Layer head left | 6,5* | 0,7 | 3,9 | 0,5 | 1,4 |
| | molecular Layer HP head right | 4,4* | 3,0 | 3,6 | 0,0 | 0,0 |
| | molecular Layer body left | 9,5** | 1,9 | 4,8* | 0,0 | 2,5 |
| | molecular Layer HP body right | 10,9*** | 2,5 | 3,5 | 0,0 | 0,5 |
| | gcmldg head left | 6,9** | 0,9 | 3,4 | 0,5 | 1,6 |
| | gcmldg body left | 5,9* | 1,7 | 5,3* | 0,0 | 1,1 |
| | gcmldg body right | 6,4* | 2,4 | 5,1* | 0,0 | 0,0 |
| | tail left | 4,0* | 0,5 | 4,7* | 0,0 | 0,9 |
| | tail right | 6,3* | 0,8 | 3,7 | 0,0 | 0,3 |
| **Amygdala** | whole left | 2,7 | 0,3 | 5,1* | 0,0 | 0,1 |
| | accessory basal nucleus left | 5,6* | 0,5 | 5,5* | 0,0 | 0,5 |
| | accessory basal nucleus right | 7,0** | 4,8* | 2,7 | 0,0 | 0,0 |
| | anterior amygdaloid area left | 0,0 | 0,0 | 6,3* | 0,0 | 0,0 |
| | anterior amygdaloid area right | 4,0* | 3,2 | 5,1* | 0,0 | 0,0 |
| | basal nucleus right | 2,9 | 3,9* | 1,6 | 1,6 | 0,1 |
| | central nucleus left | 6,4* | 0,0 | 0,0 | 0,0 | 0,0 |
| | central nucleus right | 4,8* | 3,6 | 0,0 | 0,0 | 0,0 |
| | corticoamygdaloid transition left | 0,8 | 0,0 | 6,3* | 0,8 | 0,0 |
| | lateral nucleus left | 2,1 | 0,1 | 4,6* | 0,0 | 0,1 |
| | paralaminar nucleus left | 0,0 | 0,0 | 0,0 | 0,0 | 0,0 |

Legend: *p≤.05; **p≤.01; ***p≤.001; agi = agitation, app = appetite changes, F = statistical value from the ANCOVA Model, Left = left hemisphere, right = right hemisphere, CA = Cornu Ammonis, gcmldg = granule cell layers of the dentate gyrus.

**Supplementary table 4.** Corrected ANCOVA model for differences between CN-non-converted vs CN-converted.

| | | **Simple effect** |
|---|---|---|
| **Domain** | Variable | F-Values |
| **Cognitive** | $2^{nd}$ recall of RAVLT | 4,4* |
| | Digit Span Backward | 2,5 |
| | Semantic Lexical Evocation "Vegetables" | 4,5* |
| **Cingulate** | posterior ventral gyrus right | 5,0* |
| **Frontal** | inferior orbital gyrus right | 3,9* |
| **Temporal** | middle gyrus right | 5,2* |
| | pole gyrus right | 4,5* |
| **Hippocampus** | whole right | 7,0** |
| | body right | 8,1** |
| | presubiculum body right | 8,1** |
| | subiculum head left | 8,3** |
| | subiculum body right | 10,4** |
| | ca1 body left | 5,8* |
| | molecular Layer body right | 4,0* |
| | tail left | 6,3* |
| | tail right | 7,4** |
| **Amygdala** | anterior amygdaloid area left | 7,8** |
| | corticoamygdaloid transition right | 7,2** |
| **Thalamus** | whole left | 5,8* |
| | central lateral left | 2,0 |
| | paratenial right | 0,0 |
| | pulvinar anterior left | 8,9** |
| | pulvinar medial left | 11,4*** |

Legend: *p≤.05; **p≤.01; ***p≤.001; F = statistical value from the ANCOVA Model, Left = left hemisphere, right = right hemisphere, CA = Cornu Ammonis
